# BMI Interacts with the Genome to Regulate Gene Expression Globally, with Emphasis in the Brain and Gut

**DOI:** 10.1101/2024.11.26.24317923

**Authors:** Rebecca Signer, Carina Seah, Hannah Young, Kayla Retallick-Townsley, Agathe De Pins, Alanna Cote, Seoyeon Lee, Meng Jia, Jessica Johnson, Keira J. A. Johnston, Jiayi Xu, Kristen J. Brennand, Laura M. Huckins

## Abstract

Genome-wide association studies identify common genomic variants associated with disease across a population. Individual environmental effects are often not included, despite evidence that environment mediates genomic regulation of higher order biology. Body mass index (BMI) is associated with complex disorders across clinical specialties, yet has not been modeled as a genomic environment. Here, we tested for expression quantitative trait (eQTL) loci that contextually regulate gene expression across the BMI spectrum using an interaction approach. We parsed the impact of cell type, enhancer interactions, and created novel BMI-dynamic gene expression predictor models. We found that BMI main effects associated with endocrine gene expression, while interactive variant-by-BMI effects impacted gene expression in the brain and gut. Cortical BMI-dynamic loci were experimentally dysregulated by inflammatory cytokines in an *in vitro* system. Using BMI-dynamic models, we identify novel genes in nitric oxide signaling pathways in the nucleus accumbens significantly associated with depression and smoking. While neither genetics nor BMI are sufficient as standalone measures to capture the complexity of downstream cellular consequences, including environment powers disease gene discovery.

## Introduction

The utilization of genetics in medicine has drastically expanded over the last 30 years through the identification of a growing body of rare and common genetic variation that contributes to diseases across specialties. While high penetrance rare variation can be tested in the clinic and is relevant to the clinical management of a subset of mendelian cases^1,2^, but the clinical translation of common variation, which cumulatively represents the bulk of heritability in complex traits, has proven more challenging^3,4^. Much success in identifying trait-associated common variants has come from genome-wide association studies (GWAS). This population-level approach maximizes power to identify disease-associated single nucleotide polymorphisms (SNPs)^5^, yielding hundreds of genome-wide significant loci across a very large number of complex traits. However, discovery of causal variants from GWAS is challenging; this difficulty stems from the complexity in pinpointing causal alleles from complex LD structures, translating population-level statistics to individual-level associations and, on a larger scale, because many of the common diseases in the clinic are multifactorial, i.e. due to a combination of genomic and environmental factors^6^. Moreover, GWAS will miss SNPs that are associated in only certain environmental contexts or SNPs with varying gene-by-environment (GxE) effect sizes^7^. Dynamic variant effects may be a core reason, in addition to ancestry and diagnostic bias, why population-level predictors derived from GWAS (e.g., polygenic risk scores) perform poorly on an individual level and have not successfully been translated to patient care. To truly capture the predictors of multifactorial disease development, we need to consider both genetics and environment in disease risk models.

A global finding from GWAS shows disorder risk-increasing SNPs are enriched for expression quantitative trait loci (eQTL) - variants with the functional capacity to regulate gene expression^8^. This enrichment of eQTLs may imply that the primary modality by which GWAS variants affect disease is through a cumulative impact of gene expression in disease-relevant tissues. Understanding eQTL architecture, and integrating these with GWAS summary statistics, yields biological insights and interpretability, for example allowing researchers to implicate a gene, tissue or cell type involved in the complex trait. However to-date, studies of eQTLs have focused on static associations despite evidence that eQTL activity may be highly context-specific, with dynamic effects sizes across tissues, cell-types, sex, hormonal state, developmental stage, and immune activation^9,10,11,12,13,14^, with implications for risk-estimates of common immunologic disorders and cell types^10,11^.

We propose that studies that integrate only static eQTL estimates fail to capture important context-specific associations. By characterizing and incorporating dynamic estimates into eQTL architecture, and integrating these estimates with GWAS, we hope to move beyond static estimates and into a personalized understanding of dynamic risk. Here, we focus on body mass index (BMI), a key physiological context with potential relevance to many complex traits. Obesity, often defined as BMI of >30, is rising across the globe^15^, with important physiologic impacts including inflammation, oxidative stress, and hormonal dysregulation^16,17,18^. The combined morbidity of obesity-associated cardiometabolic, oncologic, psychiatric, and neurologic conditions is substantial^,19,20,21^. Many efforts have been undertaken to use GWAS to identify factors associated with genomic susceptibility to these complex disorders ^22,23,24^, yet no studies have evaluated the impact of the environment of obesity on genomic risk estimates. In this study we test whether BMI (body mass index: kg/m^2^), here used as a proxy for the endocrinologic physiologic environment, interacts with the genome to regulate gene expression across tissues. We aim to identify BMI-dynamic genomic variants, postulate cell-type specific regulatory BMI-dynamic mechanisms, and identify novel genes and pathways relevant to complex disease, which are missed with conventional GWAS approaches.

Previous work has shown that cell type proportion may be a key mediator in eQTL and environmental interactions^25^ and is a vital consideration in defining gene-by-environment findings. To parse the impact of cell type on our BMI-dynamic eQTL effects, we perform cell type deconvolution across body systems and identify novel associations of BMI and cell fraction. We postulate cell-type specific dynamic mechanisms using transcription factor binding affinity prediction and human induced pluripotent stem cell (hiPSC)-derived glutamatergic neurons treated with pro-inflammatory factors. We build gene expression predictor models to predict tissue-specific gene expression from BMI and SNP interactions and discover novel genes that have a BMI-dynamic relationship with brain traits in a large-scale biobank.

## Results

### Defining the Impact of BMI in the regulation of Gene Expression

We implemented an eQTL linear regression framework to test for the main and interactive effects of SNPs and BMI on gene expression (**Fig 1A**). First, we followed the classic eQTL approach to identify the main effects of SNPs on expression without regard for environmental context; we term these **Base-eQTL** and **Base-eGenes**. The number of Base-eGenes was highest in the GTEx tissues with the largest sample size (cor=0.89), in line with the published GTEx v8 cis-eQTL analysis^26^ (**Fig 1B**). To determine the impact of BMI directly on gene expression, we tested for differentially expressed genes in the context of BMI **(BMI DE-genes, Fig. 1C,1D).** We find BMI-DE genes in 18 non-brain tissues, with most in the endocrine system including subcutaneous adipose (n=2341 genes), skeletal muscle (n=1894 genes), and skin-not sun exposed (n=1823 genes) (**Fig 1D**). We identified 7,563 total BMI DE-genes (FDR < 0.05), 4,199 of which are negatively associated with BMI and 3,364 are positively associated (**Fig 1C**), including known BMI-associated genes such as *LEP*^27^, *SLC27A2*^28^, and *GPD1L*^29^.

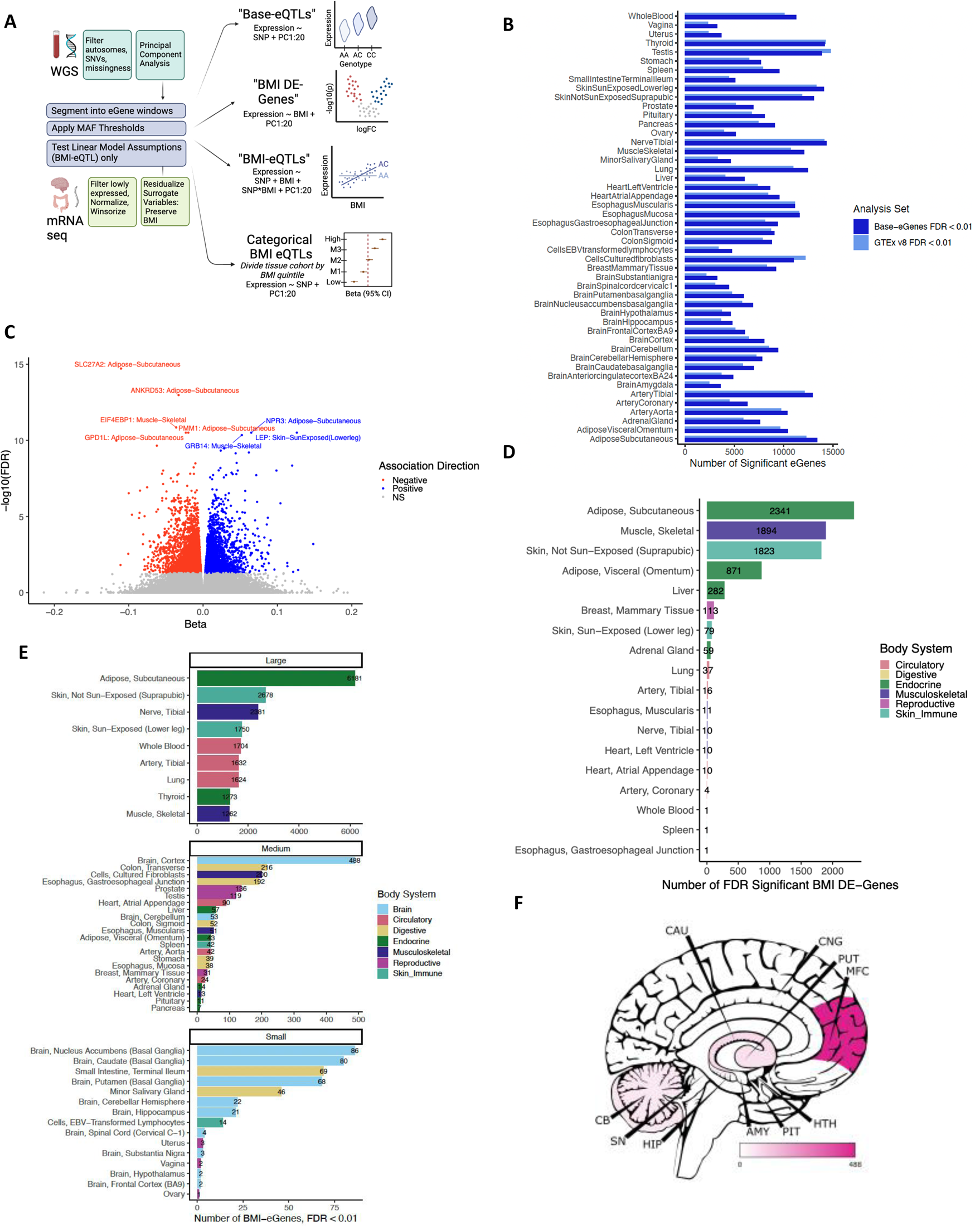

We next examined whether BMI impacts gene expression indirectly through interactions with SNPs, called **BMI-eQTL,** and their associated eGenes, called **BMI-eGenes**, by adding an interaction term to the classic framework (*Exp ∼ SNP*BMI + SNP + BMI + PC1:20).* Because eQTL discovery is correlated with sample size, we applied QC specific to the large (n≥500), medium (500>n≥200), and small tissues (n<200). Among large tissues, subcutaneous adipose and non-sun-exposed skin contained the most BMI-eGenes. Among the mid-size tissues, brain cortex and transverse colon had the most BMI-eGenes, and within the small tissues nucleus accumbens (NAc), caudate in basal ganglia and small intestine had the most BMI-eGenes (**Fig 1E**,**1F**). A portion of BMI-eGenes overlap both Base-eGenes and BMI DE-genes in subcutaneous adipose, however in cortex the majority of BMI-eGenes are not Base-eGenes (**Supplement Fig 2A-B**). Base-eGenes have greater overlap across tissues compared to BMI- eGenes (Average Szymkiewicz–Simpson (SS) overlap coefficient: Base-eGenes SS=0.67; BMI-Genes =0.066; **Supplement Fig 2C-D**).

### Characterizing BMI-eGenes Function and Interaction Direction

Base-eGenes are generally tolerant to loss-of-function variation and have limited constraint in the genome, as opposed to GWAS hits which are more constrained^30^. We assessed whether BMI-eGenes show a similar pattern by annotating the probability of loss-of-function intolerance (pLI) score for our eQTL sets and testing for enrichment of high pLI scores in Base- and BMI-eGenes compared to background. We found that BMI-eGenes are enriched for loss-of-function intolerance in subcutaneous adipose (p=7.5×10^-25^), brain cortex (p=0.034), and cerebellar hemisphere (p=0.01) (**Fig. 2A**), while Base-eGenes have no enrichment of constrained genes. BMI-DE genes are enriched for constrained genes in subcutaneous adipose (p=1.9×10^10^), skeletal muscle (p=7.6×10^-7^), liver (p=4.4×10^-2^), and skin not sun exposed (p=6.9×10^−4^).

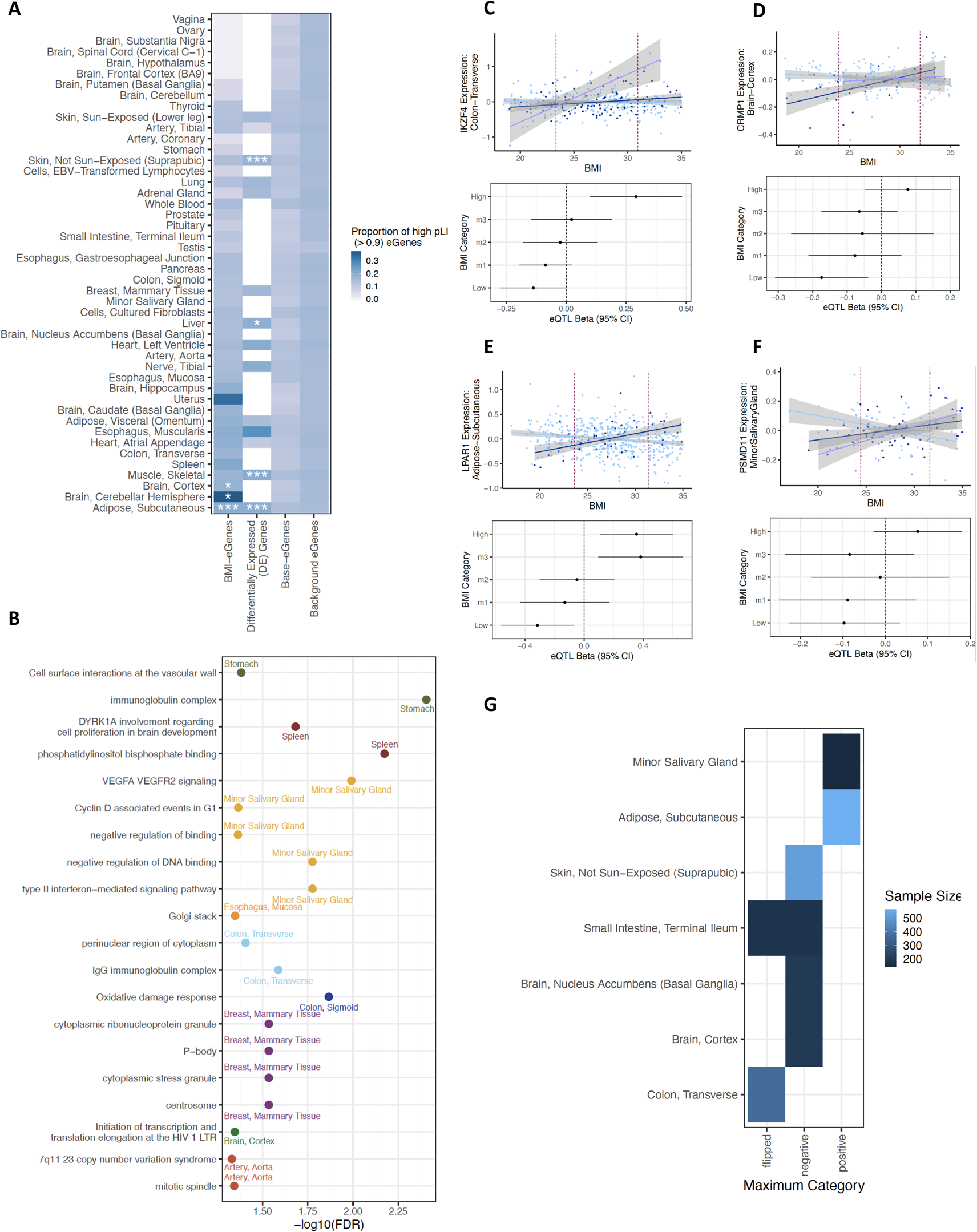

Gene set enrichment reveals that the Immunoglobulin complex is interactively regulated by SNPs and BMI in the stomach (p=3.9×10^-3^) and transverse colon (p=0.026). Cancer pathways are enriched across tissues, including cyclin D - associated events in G1 in minor salivary gland (p=0.043) and P-body in breast tissue (p=0.029), and oxidative damage response on colon-sigmoid (p=0.014) (**Fig. 2B**).

For subsequent analyses, we selected a subset of tissues across the sample size categories that had the most BMI-eGenes, including adipose subcutaneous and skin not sun exposed in the large category, transverse colon and brain cortex in the medium, and small intestine and nucleus accumbens (NAc) in the small. We added minor salivary gland due to the presence of numerous gene set enrichments and in total have seven tissues prioritized for future study.

Thus far, we have analyzed the properties of the significant eGenes identified in the eQTL analyses. To characterize the number of independent genomic loci in each cis region that regulate each eGene, we performed stepwise conditional analysis. We selected the independent SNPs from conditional analysis, called the top SNP(s), and identified SNPs in LD with the top SNPs to generate eQTL credible sets for each locus in the Base-eQTL and BMI- eQTL analyses. We find 10,451 credible sets of BMI-eQTL across the 7-tissue subset, including 9,612 unique SNPs and 8,434 unique genes. Across tissues, Base-eGenes had more independent credible sets in the cis-region locus than BMI-eGenes, but there was a shared pattern of distribution across tissues (**Sup Fig 2E**).

To determine whether BMI*SNP interactions have a predominant direction, i.e. if high BMI is most often associated with a reduction of eQTL activity, we characterized the directionality of the independent BMI*SNP relationships. Using a decision tree based on categorical BMI models, we identified four possible directions: positive (absolute eQTL activity, i.e. beta, is larger at high BMIs), negative (absolute eQTL activity is larger at low BMI), flipped (eQTL activity is significant in high and low in opposite direction) (**Fig. 2C-2F)**, and uncertain (**Fig 1A, Supplemental Figure 3A).** Across the seven tissues, 4,167 BMI-eQTL are positive, 2,471 are flipped, 2,427 are negative, and 1,383 are uncertain. We found that the distribution of directions among BMI-eQTL were not shared across tissues. The negative direction was the largest category in cortex, NAc, skin, and small intestine while the positive direction was the largest category in subcutaneous adipose and salivary gland. We noted each tissue had BMI- eQTL in all possible directions (**Fig. 2G, Supp Fig 3A-B**).

### Parsing the Roles of Sequence and Cell Type in BMI Dynamic Processes

We next asked whether BMI interacts with the genome at certain transcription factor (TF) binding sites by comparing TF binding affinity prediction between significant Base-eQTL and BMI-eQTL. There are 67 total nominal, and 53 FDR TFs significantly enriched in BMI-eQTL compared to Base (**Fig. 3A**). The majority of TFBS (37/53) are significant in one tissue, five TFs are significant in two tissues, and two TFs, NFATC2 and NR4A2 are FDR significant in three tissues. Of note, FOXO3 in nominally significant in the most (5/7) tissues.

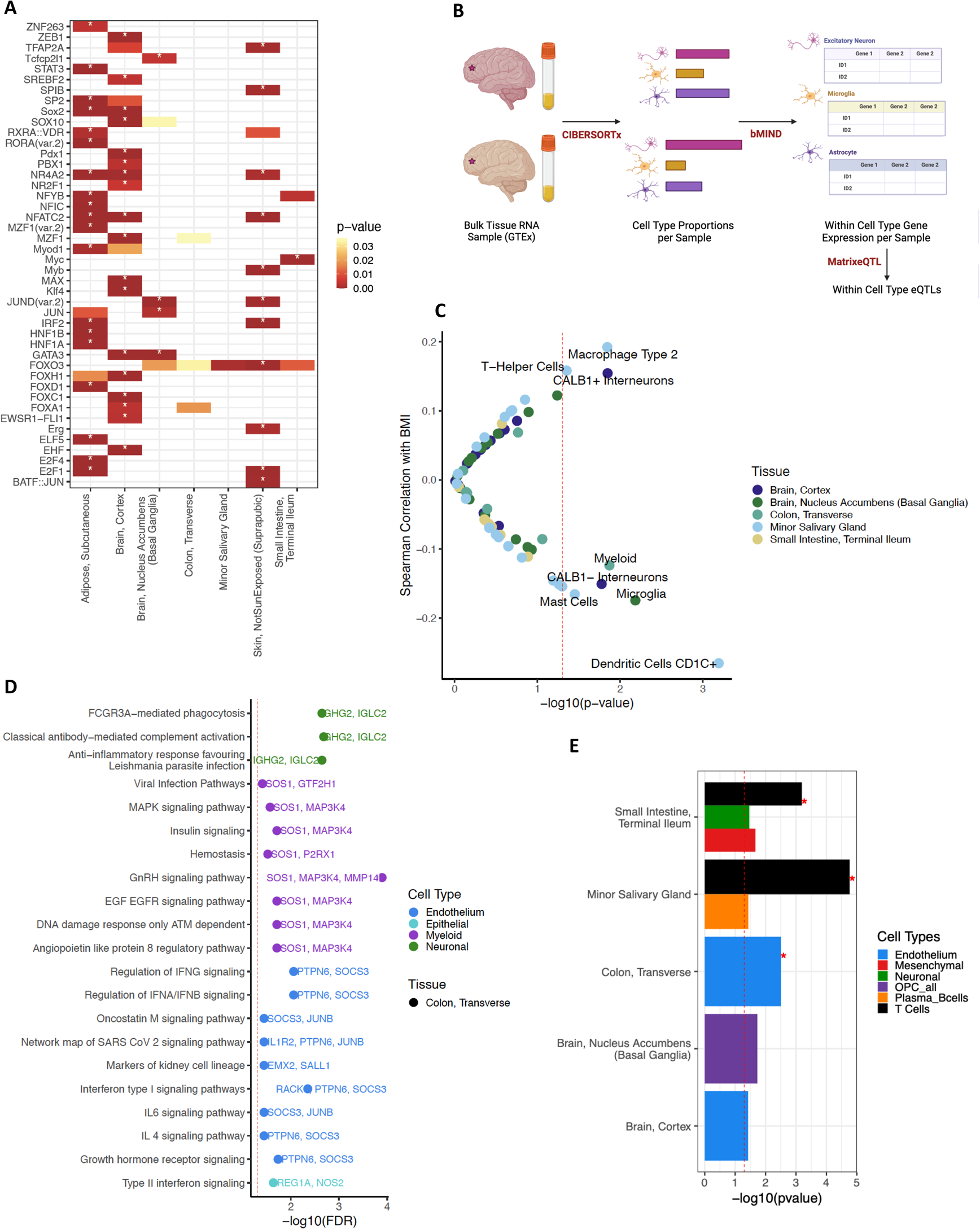

Our study uses bulk RNA sequencing data, including a combination of mRNA from all cell types in the sample. To determine whether cell type proportion plays a role in BMI-eGene discovery, we applied cell type deconvolution techniques (**Fig. 3B**). The association of BMI and cell fraction has been previously characterized in the adipose and endocrine system^31^, therefore we focused on (novel) brain, gut, and salivary gland.

BMI was significantly correlated with estimated cell type fractions of four immune cell types in salivary gland: T-helper cells (cor=0.16, p=0.044), macrophage Type 2 (cor=0.19, p=0.014), mast cells (cor=-0.16, p=0.035), and dendritic cells type 2 (cor=-0.26, p=6.4×10^-4^) (**Fig. 3C**). The positive association between BMI and macrophage is supported by analyses completed in adipose^31^. BMI was negatively correlated with estimated proportion of microglia in NAc (cor=-0.17, p=6.6×10^-3^) and with myeloid cells in the transverse colon (cor=-0.12, p=0.013). To determine the impact of cell fraction on BMI-eGene discovery, we repeated the interaction eQTL analysis correcting for cell fraction. A range of 33-65% of BMI-eGenes remain significant after cell type correction (Supplemental Figure 4A). Our data indicate that both immune cell type proportion and cis regulatory motifs underline the BMI-dynamic signal identified.

We identify enrichment of cell-type specific gene sets in genes significant after cell type correction in transverse colon (Fig 3E) and cortex (Supp). In colon, we find antibody-mediated complement activation is enriched in neuronal cells (p_FDR_ =2.1×10^-3^), while myeloid cells have enrichment for insulin (p_FDR_ =0.019) and GnRH (p_FDR_ =0.018) signaling, among others. In endothelium, inflammatory factors including IL-6 (p_FDR_ =0.036), IL-4 (p_FDR_ =0.036), and INFA/INFB (p_FDR_ =8.7×10^-3^) are enriched, while in the epithelial cells type II interferon (p_FDR_ =0.023) is enriched. To test for cell types enriched with BMI-dynamic effects, we tested for differential enrichment of lead BMI-eQTL compared to Base-eQTL in specific cell types. We found that BMI-eQTLs were significantly enriched in T-cells in minor salivary gland (p_FDR_ =6.5×10^-4^) and small intestine (p_FDR_=0.012), in endothelium in transverse colon (p_FDR_ =0.039) and nominally enriched in endothelium in cortex (p=0.037) (**Fig 3E**).

### Deciphering Mechanisms of BMI Dynamic Effects using *in Vitro* Models

Because BMI-dynamic processes are in part inflammatory, one possibility is that overactivated microglia may release proinflammatory cytokines that crosstalk with neurons ^32,33,34^]. To empirically resolve whether inflammatory signals indeed mediate cortical BMI-eQTL activity in neurons, we conducted ATACseq on hiPSC-derived *NGN2*-induced glutamatergic neurons at baseline and in the context of three acutely treated (48 hours) pro-inflammatory cytokines IL-6 (60 ng/uL), INFa-2b (500 UI/mL), and TNFa (100ng/mL)^35^. An adapted activity-by-contact (ABC) model using STARE^36^ predicted dynamic enhancer-gene interactions across inflammatory contexts, linking BMI-eQTL to predicted target genes (**Fig 4A**). While only 37% (256/690) of base-eQTL top SNPs were active at baseline (ABC >= 0.1), 73% (141/194) of BMI- eQTL top SNPs were active at baseline. In most cases, BMI-eQTL top SNPs in the credible set were the SNPs with the maximum ABC score (max-ABC) for that gene (81%, 131/161), but this was not true from base-eQTLs, which were more likely to have a max-ABC that was in LD with the top SNP (33% 230/689).

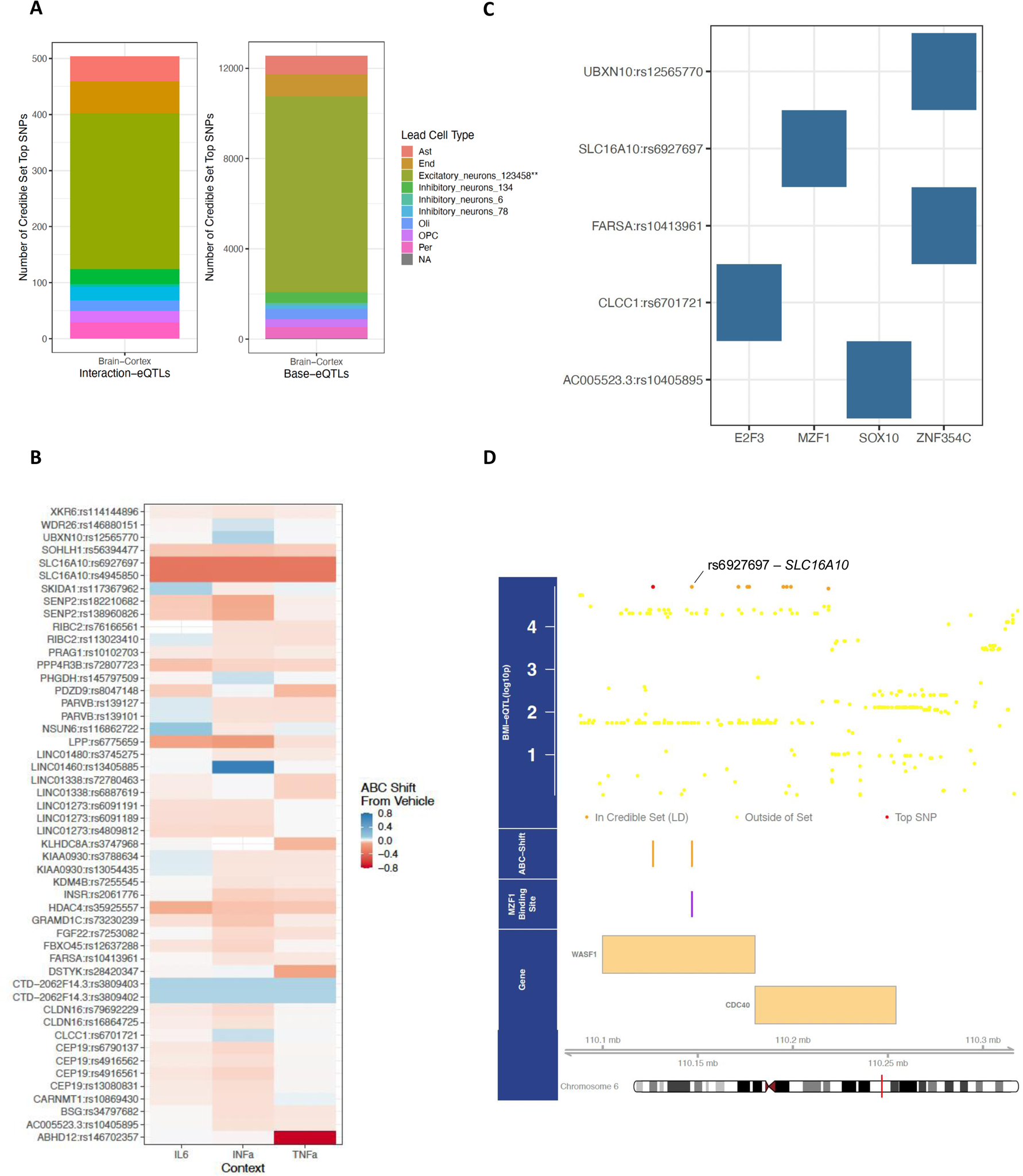

In all BMI-eQTL sets, we identified 50 SNPs regulating 37 genes that have an ABC-score shift from vehicle >= 0.02 for at least one context (**Fig 4B**). INFa shifted the most BMI genes(n=42), while TNFa the least (n=23). Regulatory shifts for eQTL activity could be shared or distinct across contexts; for example, *ABHD12*, displayed a TNFa-specific shift. TF-binding disruption is predicted to underlie five of the genes identified (**Fig 4C**), with altered ZNF354C binding predicted to distupt two genes *UBXN10* and *FARSA* found on different chromosomes. *SLC16A10,* a monocarboxylate transporter also called MCT10, showed context-wide regulatory shifts at two SNPs (rs6927687 and rs4945850), one of which (rs6927687) had a predicted impact on MZF1 binding and so was likely the BMI-dynamic regulatory SNP of *SLC16A10* (**Fig 4D**), which had pan context-specific accessibility at its promoter region (**Sup Fig 4D**).

### Advancing Brain Trait Gene Discovery using BMI-aware Gene Expression Predictor Models

Our results show that BMI interacts with the genome to regulate gene expression. Predicting gene expression from genotype is well established and allows researchers to add gene- and tissue-level context to GWAS studies. However, available predictor models have focused on prediction only using genomic data, called genetically regulated gene expression (**GREx**). Here, we created new predictor models that incorporate BMI, allowing prediction of dynamic, context-specific gene expression. We trained predictor models that include main effects of genotype, BMI, and interaction terms, and compared our BMI-dynamic models to existing GREx predictors using PredictDB-PrediXcan^37^ (**Fig 5A)**. BMI-dynamic models significantly predicted fewer genes (N_GREx_=3670-8650, N_BMI-dynamic_=1653-5508) (**Fig 5B)**, with both shared and novel genes compared to GREx predictor models (N_novel_=262-361, **Sup Fig 5B).** For genes that have models made using both methods, BMI-dynamic models that include interaction terms explained significantly more variance in gene expression than GREx models (p_adipose_= 2.58×10^-35^, p_cortex_=5.13×10^-23^, p_NAc_ =3.08×10^−23^, p_skin_ =7.85×10^−28^, p_small intestine_=7.32×10^-19^=7, p_colon_ 4.01×10^-21^=7)(**Fig 5C**). We found that BMI is primarily included as only a main effect in endocrine tissues, including skin and subcutaneous adipose, while in the brain and gut we predominately found interactive effects and minimal BMI main effects, which mirrored the results from the BMI DE-gene and BMI-eGene analysis (**Fig 5D**).

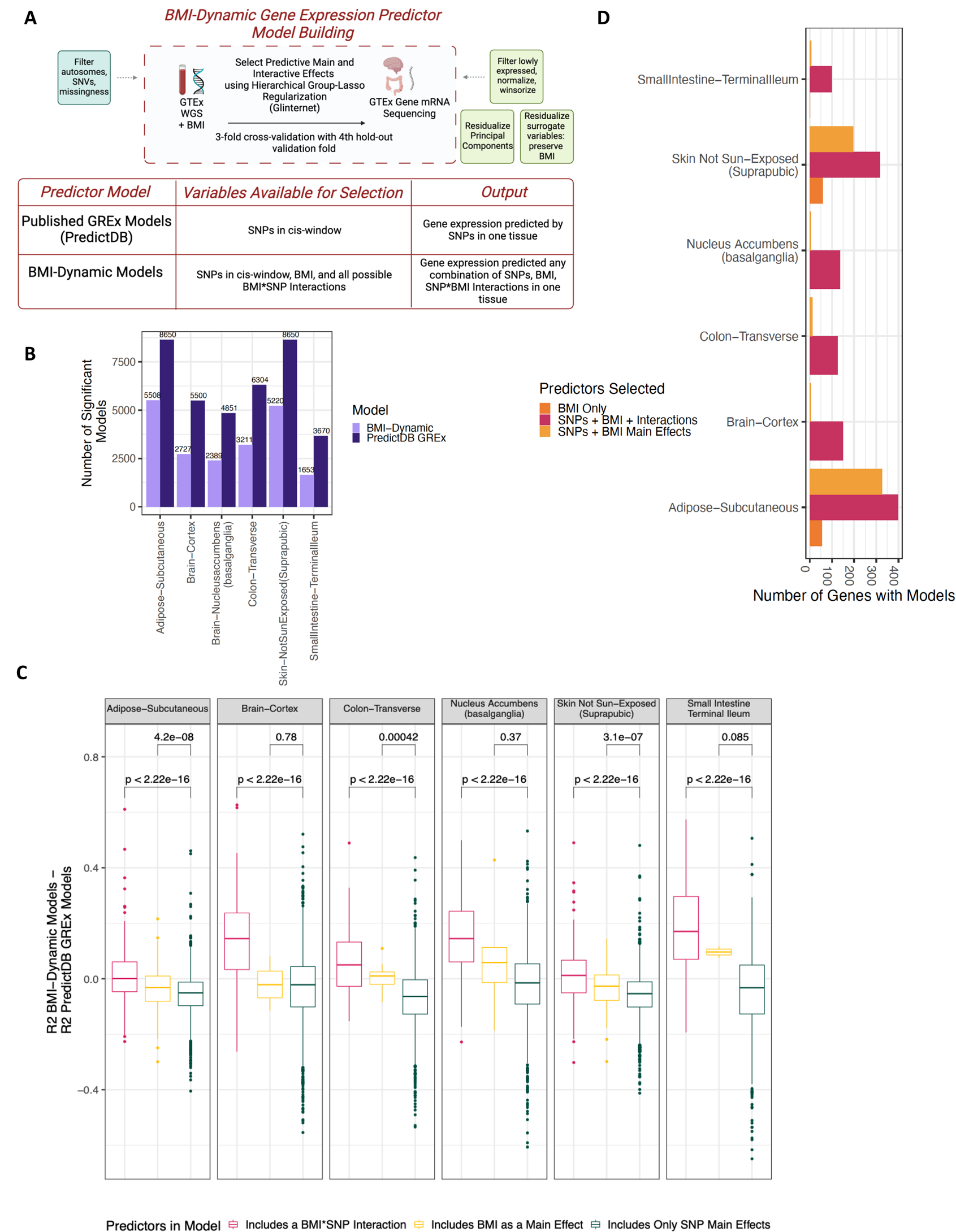

We hypothesized that by applying BMI-dynamic predictor models to brain traits, we may enhance gene discovery over methods that predict using only SNP effects. We applied our models to predict BMI-dynamic gene expression in cortex, NAc, and subcutaneous adipose for all UK Biobank participants, and tested for the association of predicted expression with depression and current smoking (**Fig 6A**). Across tissues and traits, BMI-dynamic models identified more significant associations (**Fig 6B**) compared to GREx models. Most of these additional associations occurred in genes where the BMI-dynamic predictors include BMI as a main effect with or without an interaction (**Fig 6D**). The significant gene associations in depression in NAc (p=0.011) and adipose (p=0.03) were enriched with nominally significant S- predixcan MDD-GWAS hits (**Sup Fig 6A**). In the NAc, the MHC locus is enriched in the BMI- dynamic depression association (pFDR=9.13 x10^-5^) driven by genes *HLA-G*, *HLA-DQB2*, *HLA-DMB*, and *HLA-DMA* (**Fig 6C**). We identified enrichment in the BMI-dynamic NAc association for glutathione peroxidase (pFDR=0.029) in smoking and hydrolase activity (pFDR=0.029) in smoking and depression, both components of the oxidative stress pathways. The significant BMI-dynamic trait-gene associations are not commonly shared across tissue, with more overlap across traits **(Sup Fig 6D-H).**

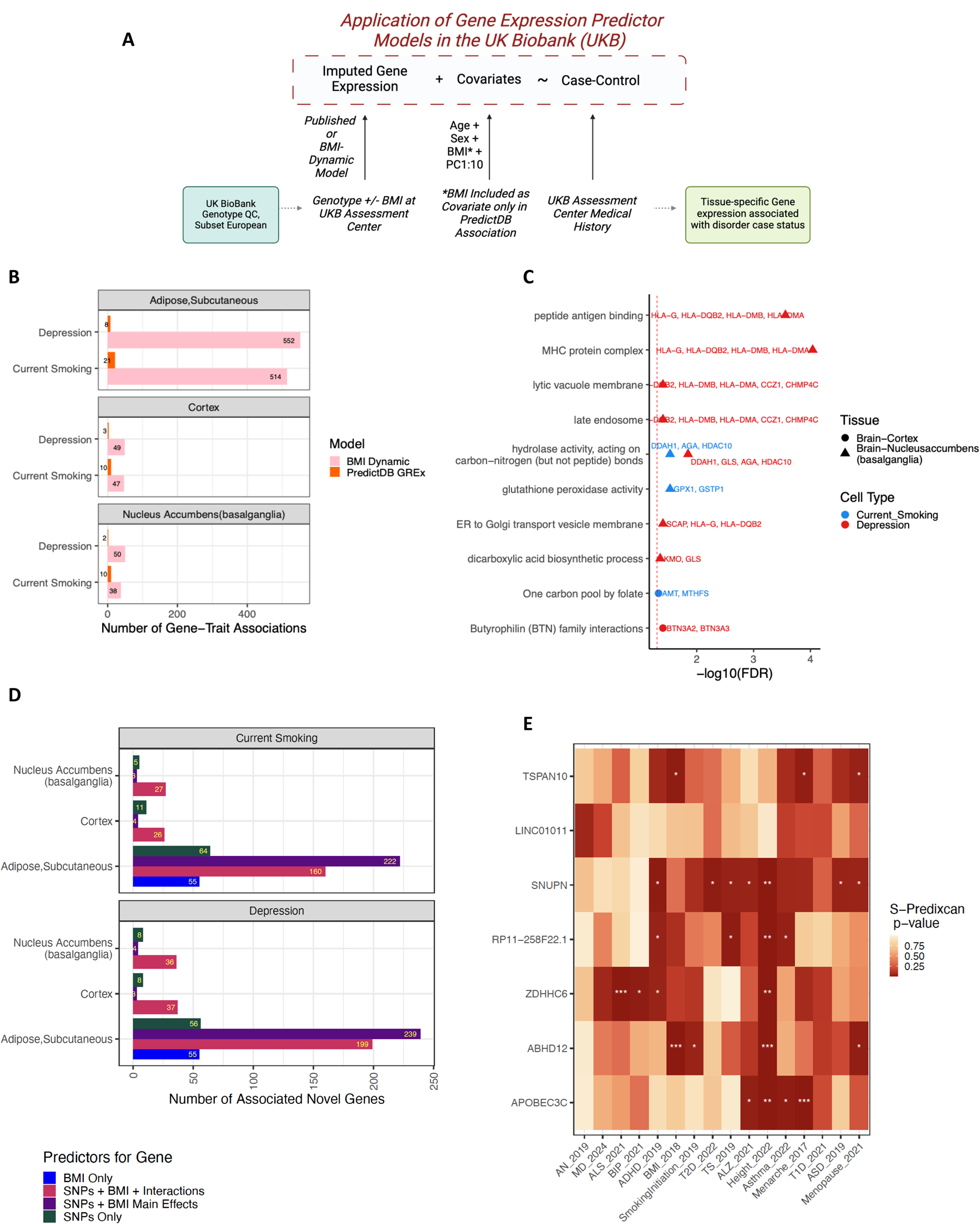

We compared the published GREx and BMI-dynamic associations in the NAc smoking associations and found a subset of the BMI-dynamic model genes only contain SNPs as predictors but are not significant in the published GREx analysis **(Sup Fig 6B-C)**. In this set, we find genes that overlap our BMI-eGene analysis and iPSC analysis, including *ABHD12*, indicating they may be BMI-dynamic while not including BMI as a main or interactive effect in the model. We surmise that these genes have BMI effects of expression which have been preserved in our BMI-specific RNA sequencing quality control but were not in the BMI-agnostic QC from previous studies.

We found 215 genes in adipose and nine genes in cortex significant in the BMI-dynamic depression and smoking associations that were also significant BMI-eGenes. To identify phenotypic associations beyond depression and smoking, we tested for the nominal association across GWAS traits using S-Predixcan in the nine cortex overlap set (**Fig 6E**). Of the nine genes, seven have PredictDB-GREx models published and can be used with the S-Predixcan tool. Of note, this analysis will not capture the true breadth in association, as S-Predixcan is based on models that do not learn BMI effects, but it gives insight toward nominal associations that might be stronger when BMI is considered. We see most of the genes (6/7) are nominally significant in the height or BMI GWAS, indicating these SNPs might serve as both a predictor for BMI and have dynamic regulatory action in the context of BMI. The gene *SNUPN* has the most associations across traits including attention-deficient hyperactivity disorder, type 2 diabetes, Tourette syndrome, Alzheimer’s disease, height, autism spectrum disorders, and age at menopause.

## Discussion

In this work we demonstrate that BMI interacts with the genome to (dys)regulate gene expression within and beyond the endocrine system. Gene-by-BMI effects on gene expression in the brain are most pronounced in the reward regions including the basal ganglia and cortex, but not in classical obesity-associated regions such as the hypothalamus, which had the third lowest number of genes regulated by BMI*SNP interactions across tissues. The observation that the striatum has more BMI-dynamic genetic regulation than the hypothalamus expands our understanding of the variety of brain regions involved in metabolic response. We note that we did not identify any FDR significant BMI-differentially expressed genes in the brain; instead, BMI DE-genes were primarily found in endocrine tissues. This is intuitive as the endocrine system is responsible for adapting to the fed or fasted state while the brain is protected from direct BMI effects. Adipose and cortical BMI-eGenes were enriched for genes that are highly constrained, and thus more vital for reproductive fitness. Previous work has shown that classic eQTL are generally less constrained while GWAS hits demonstrate greater constraint^30^ which constitutes a core difference between eQTL and GWAS that limits colocalization and the mapping of common eQTL to disease trait. We show here that context-specific eQTL look more like GWAS hits in terms of loss-of-function intolerance and may be more likely to be trait-associated.

The epidemiological associations of BMI with adverse outcomes such as all-cause mortality exhibit a U-shaped pattern of effect, where mortality is increased at low and high ends of the BMI-spectrum^38^. To identify eQTL effect differences at high versus low BMI, we implemented a directionality analysis of BMI-eQTL using categorical BMI eQTL models. We find that BMI-eQTL can exhibit multiple patterns, where some eQTL are more active at high and others at low, and there is not an apparent pattern across tissues. In the adipose, BMI-eQTL are most often active at high BMI whereas in the brain most are more active at low BMI, but eQTL in both directions exist in both. This highlights the pattern of BMI-dynamic effects described, some people may have more eQTL active, and be more susceptible, at high BMI and others at low. Further work about context-wide susceptibility scores is needed to precisely sum the effects of trait risk in the context of environmental factors.

We used the publicly available dataset GTEx to explore these relationships, however GTEx is a postmortem dataset and does not represent the full range of BMI, especially the very low and very high BMI ranges. Additionally, the associations of BMI and health outcomes are reported to be related to sex, age, and developmental stage ^39,40^, which are not represented in just one dataset and may impact replication when applying to other datasets with different demographic features. Further research into the timing of BMI-SNP relationships in the context of age and sex are vital to parsing the impacts of the interactions identified above.

Previous literature has shown that obesity impacts cell type proportion in adipose tissue, where macrophage infiltration and abundance increases in the context of obesity^41^, and in the brain where volume loss and migroglial activation are noted^42,43^. We see a similar pattern of correlation between microglia and BMI in the nucleus accumbens, however the correlation is negative. Previous reports have shown that over-activated microglia are targets of apoptosis^44^, which may be an explanation for the negative relationship. We found that the surrogate variables used to residualize the expression matrix correlated with cell fraction and therefore are corrected for in our BMI-eQTL equation-however we preserved the effects of BMI in the residualization and therefore a portion of our signal may still be due to BMI effects on cell type that are difficult to deconvolute. This is a limitation of the study and should be addressed in single cell datasets with BMI as a measure in donors to validate our findings.

We find evidence that the expression of genes in inflammatory pathways such as IL-6 and INFA/B/G are interactively regulated by SNPs and BMI in the endothelium of the transverse colon. The literature reports IL-6, TNFa, and interferons^45,46^ (among other inflammatory factors) underlie the physiologic effects of obesity. The endothelium has been attributed to both response to and production of immune responses^47^, and this work highlights the role of the colonic endothelium as a BMI-dynamic pro-inflammatory cytokine producer, which may be a vital aspect of inflammatory gut disorders.

We hypothesize that, in the brain, overactive microglia secrete pro-inflammatory cytokines^48,49,50^ which cross-talk with neurons^51,32^, and induce the BMI-dynamic loci identified in this study. We tested this hypothesis by washing a glutamatergic iPSC neuronal system with pro-inflammatory cytokines, measuring baseline and context-specific chromatic accessibility with ATACseq in neurons, and testing for overlapping effects with BMI-eQTL using an activity-by-contact model. We find evidence that pro-inflammatory cytokine signals mediate context-specific regulation for several BMI-eGenes in shared and distinct cytokine patterns. Surprisingly, we see the most context-specific shits in our data in the context of interferon-alpha, which is less often cited as a player in obesity than TNFa and IL-6. Further work on the interrogation of interferon-mediated inflammation in the CNS and nucleus accumbens, including the effects if INF-G, should be completed^52^. Additionally, iPSC systems model fetal-like cell states while the effects found here were in postmortem adult. Synchronization of developmental stages may identify even more overlap.

We have shown that the functional capacity to regulate gene expression for a subset of SNPs is not static in a population, and instead is dynamic in the context of their physiology. This has implications for modeling BMI-associated complex disease genetic associations. We have shown that by adapting genomic predictor models to include the effects of BMI, we find more disease-associated genes. We find more disease-associated genes despite a lower number of significant models compared to the gold-standard GREx approach using Predixcan-PredictDB. This may be due to differences in quality control such as a more lenient MAF threshold or the decision to use the entire ancestry cohort^53^. It also may be a byproduct of the different machine learning models used, reflected by the differing number of features selected. A final contributor is the increased variance explained in gene expression denoted by BMI-aware models, which capture additional direct and indirect effects of BMI. If the field can adapt GWAS and post-GWAS approaches including polygenic risk scores to be environment-aware and fit for the individual, we likely will see better predictive performance in the clinic.

The gene discovery powered by environment-aware modeling is not only relevant for risk prediction, but also highlights molecular pathways associated with BMI-associated disorders that may be targeted for therapeutics. We find evidence that depression is a metabo-immune disorder in the brain with enrichments in both the MHC region and various metabolic pathways. Hydrolase activity on carbon-nitrogen bonds, which is driven by signal is *DDAH1*, *AGA*, *HDAC10*, and *GLS,* was significant in both depression and smoking in the nucleus accumbens. *DDAH1* is a nitric oxide synthase regulator and knockout of this gene impacts dopamine metabolism^54^. The nitric oxide (NO) has been implicated across psychiatric disorders including depression^55,56^ and has been successfully targeted in mice for prevention of stress-induced depression^57^. Here we show that metabolism of NO is regulated jointly by SNPs and BMI in the nucleus accumbens and associated across psychiatric disorders, proving one potential mechanism of NO effect in the brain.

We see other mechanisms of oxidative stress in our analyses including the significant enrichment of glutathione peroxidase pathway, driven by *GPX1* and *GSTP1,* in the smoking analysis in NAc, oxidative damage response in BMI-eGenes in the sigmoid colon and cytoplasmic stress granule in the breast. *ABHD12,* a lipase that, when knocked out in activated microglia, have altered morphology and increased phagocytosis^58^, was significant in all three analyses: it is a BMI-eGene, that is dynamic in the context of TNFa in iPSCs and significant in the smoking TWAS in the UK Biobank in cortex and NAc. *ABHD12* expression increases in the context of an inflammatory stimulus and tempers phagocytosis to reduce oxidative stress^58^. Oxidative stress has been linked to cancers, neurologic disorders, gastrointestinal disorders, and here we show it is regulated by common genetic variants and individual environment. We are providing the BMI-dynamic gene expression models for public use to promote future studies across disorders.

From these results, we assert that individuals carrying a varying number of BMI- dynamic SNPs can have gene expression that is more dynamic, or perhaps dysregulated, in the context of a changing BMI than individuals without these BMI-dynamic SNPs. This work supports the guidance that medical counseling on BMI as an isolated measure is insufficient to capture the personalized nature of risk for obesity-associated disorders. BMI is a useful tool in that it is free and easy to measure and represents an individual’s weight change over time; but we are continually recognizing that BMI as a stand-alone is inadequate when defining individual risk or comparing across populations. Genetic effects, which can be elicited with a family history, in combination with environmental effects such as BMI changes, sex, age, and psychosocial factors need to be considered in unison while providing counseling and running genomic models related to multifactorial disorder risk.

## Methods

We analyzed transcriptome samples released in the GTEx freeze v8^26^. In brief, GTEx is a publicly available resource which includes deceased donor health questionnaire, postmortem whole genome sequencing, and transcriptome (mRNA) sequencing from one or more tissues. For genomic association testing, we required each tissue have a sample size of 100 samples, which resulted in 48 tissues included in subsequent analyses. A detailed description of biospecimen collection, analyte extraction, and GTEx-completed quality control (QC) is available in the version 8 release text and supplement.

### Subject and sample outlier removal

We plotted and visualized tissue sample attributes to remove samples with outlying RIN, end base mismatch rate, transcripts detected, alternative alignment, brain weight, and split reads. We excluded height, weight and BMI outliers (+- 3SD) as well as individuals with an amputation. We excluded four individuals who had outlying height values and one individual with an outlying weight listed. The GTEx dataset ranges in age from 21-70 years with a median age of 55 and is 67% male. Race of the cohort is reported to be 86% white, 11% black, 1% Asian with remaining contributions from Native Americans and unknown. We chose to include all ancestries in eQTL generation to maximize sample size and application^53^. The BMI range in GTEx is 17.03-35.05 kg/m^2^. The median and mean BMI in this study are 27.37 and 27.33 kg/m^2^ respectively, compared to the most recent mean BMI of 30 kg/m^2^ in the United States from 2020^59^.

### Transcriptome (RNA) sequencing processing

The GTEx transcriptome release transcripts per million (TPM) and read counts, which were aligned to human reference genome GRChg38/hg38 and quantified based on GENCODE release v26. In R v4.0.3, we retained transcripts annotated as autosomal lncRNAs, protein coding, and immune-related genes. We filtered out lowly expressed genes, requiring genes have greater than or equal to 6 reads and greater than or equal to 0.1 TPM in at least 20% of samples in each tissue. We normalized read counts between samples using TMM in the edgeR v3.32.0 package^60^. We applied a winsorize function, so that any samples deviating more than three standard deviations (sd) from the mean were set to the three sd limit. We used voom from the limma^61^ v3.46.0 package to transform the normalized count data to log2 counts per million (logCPM). In order to detect hidden sources of variation such as batch or age that might confound association analyses while still retaining the effect of the environmental variable BMI we used the surrogate variable analysis v3.30.1 package^62^. We calculated the number of surrogate variables (SVs) to create using the “be” method in the sva package. Finally, we residualized the logCPM matrix with limma to correct for the SVs. This tissue-specific residual gene expression matrix was used for subsequent linear models.

### Whole genome sequencing (WGS) processing

We began our quality control with the final v8 phased VCF file provided by GTEx, which contains 46,526,292 sites. A detailed description of QC to generate this final is available from the GTEx consortium v8 supplement. Using plink v2^63^, we retained only autosomes and removed insertion-deletion (indel), multi-allelic sites, and ambiguous SNPs. We removed variants with missingness greater than 1% (--geno 0.01) or that deviated from Hardy-Weinberg equilibrium (HWE) (--hwe 0.000001). We removed samples with greater than 1% genotype missingness (--mind 0.01). To identify the largest sources of variance in the genotype data due to factors such as population structure we generated principal components (--pca) which were later used as covariates in eQTL linear models.

### Expression quantitative trait loci mapping

Toward our goal of understanding the effects of BMI on the genetic regulation of gene expression, we first ran a BMI differential expression (DE) analysis (**Eq 1**).

#### Equation 1. BMI-DE Analysis: Expression ∼ BMI + PC1:20

For the DE analysis, we applied a genome-wide FDR correction and report FDR < 0.05. We next generated three sets of eQTLs using the R package MatrixEQTL_v2.3^64^ using a cis-eGene window distance of 1e6. We set a MAF threshold of 10% for tissues with 199 or less samples, 5% for tissues with 200-499 samples, and 1% for tissues with 500 samples and over^65^. We identified significant eGenes using a hierarchical multiple testing correction procedure by first applying a local correction using eigenMT^66^. We then selected the smallest local adjusted p-value for each eGene across the genome and applied a genome-wide Benjamini & Hochberg (BH) FDR correction^65^. The first eQTL set, “Base-eQTLs”, (**Eq 2**) are blind to BMI and identified using with the modelLINEAR function in MatrixEQTL.

#### Equation 2. Base-eQTL Analysis: Expression ∼ BMI + PC1:20

Bas-eQTLs are akin to the bulk cis-eQTLs called the seminal GTEx v8 manuscript, with slight differences in QC and significant threshold assignment. The second set of eQTLs are “Interaction-eQTLs” and generated by adding an interaction term to the base model (**Eq 3**) with the modelLINEAR_CROSS function.

#### Equation 3. Expression ∼ SNP + BMI + SNP*BMI + PC1:20

We assessed significance of the beta coefficient of the interaction using the multiple testing procedure outlined above. Genes with at least one significant SNP are referred to as BMI-eGenes. Lastly, we calculated a “categorical-eQTL” set by subsetting the within-tissue cohort into BMI quintiles and calculating Base-eQTLs (**Eq 2**) within quintile. We identify significant BMI-eGenes using the coefficients of the interaction models and use the categorical models to annotate the directionality of the significant interactions.

### Testing assumptions of OLS in the significant eQTL models

We next sought to determine if the significant BMI-eQTL models met the assumptions of ordinary least squares (OLS) regression as often as Base-eQTL and were robust significant associations. There are four assumptions of OLS: linearity of the predictor and response, independence, normality of the errors, and homoscedasticity (constant error variance). The relationships of SNP and expression are assumed to be linear, as is the case in the eQTL literature. Additionally, relationships of BMI and SNP on expression were visualized and are depicted in Figure 2 and in the supplement. Independence also is satisfied as the data is not time-series. We statistically tested the assumption of normality for each model using the Shapiro-Wilk test, shapiro.test() in stats base R. We assessed the assumption of homoscedasticity using the Koenker’s studentized version of the Breusch-Pagan test, bptest() in the lmtest package, v 0.9-38.

### Replication of BMI-eQTL

We assess replication in two brain tissues using the Common Mind Consortium (CMC)^67^ Dorsolateral prefrontal cortex (DLPFC) from CMC release 4. Quality control for the CMC datasets mirrored the QC performed for the GTEx data set. One subject with XXY chromosomes and samples with outlying height, weight, and BMI values were removed from subsequent analyses. Subjects of European descent were selected for testing replication. We ran ancestry analyses using the 1000 genomes reference panel and the unsupervised analysis in the admixture tool^68^. For RNA sequencing, genes with >= 6 reads in >= 20% of samples are retained and mapped genes were normalized between samples using edgeR. Outliers were winsorized and transformed to log2CPM. We applied surrogate variable analysis to the RNA sequencing data to generate SV’s using the “be” method that preserve the effect of BMI and residualized the expression matrices with the SVs.

In contrast to GTEx and WGS data, CMC genetic data is imputed genotype calls to the HRC platform. For genotype QC, we filtered for imputation quality, Rsq >= 0.3 ^69^, removed sex mismatch, and ran relatedness in plink 1.9, removing a related sample if IBD > 0.125. In plink2, we applied missingness flags -geno 0.01, mind 0.01, and HWE 1×10^-6^ and output data in both dosage and hardcall and tested both for replication. We computed principal components in plink for later use in the eQTL model as covariates. We called Base-eQTLs and BMI-eQTLs in CMC according to the same equations as in GTEx. We assessed replication using the πι statistic from the qvalue package.

### Loss of Function Intolerance Enrichment

To test for enrichment of functionally relevant genes in our sets, we obtained the probability of loss-of-function intolerance (pLI) score from gnomAD/2.1.1^70^ for Base-eGenes, BMI-eGenes, BMI-DE genes. We assessed for enrichment for high pLI (pLI > 0.90) genes in each category compared to the background of genes using a binomial test where x= number of eGenes that are high pLI and significant, n=number of significant genes, and p= number of high pLI genes in this tissue/all genes in background of tissue.

### Gene Set Enrichment

To define enriched gene sets in the sets of BMI-dynamic loci identified, we implemented gene set enrichment using the gprofiler2 package^71^ with the ‘fdr’ correction method and a custom background set defined as all genes expressed in the tissue of interest. We tested GO, KEGG, REACTOME and Wikipathways.

### Conditional Analysis

To probe the number of independent signals in each eGene loci we employed stepwise conditional analysis in R for the Base-eGenes and BMI-eGenes. For each eGene, we generated a linkage disequilibrium (LD) matrix using the genotype data from the eQTL analysis and the cor() function in R with the pairwise complete observation setting. We identified the top SNP signal, which is the SNP with the smallest SNP coefficient p-value and all SNPs with r>0.999 correlation. We added the most significant SNP in the model and re-ran the eQTL model for the eGene window SNPs. If the SNP coefficient p-value of new most significant SNP in the iteration was less than 0.001, it was added into the model and a subsequent iteration was ran. The conditional analysis iterated until the top p-value was >= 0.001.

For the interaction conditional analysis, we repeated this process except at start of the conditioning we add the top SNP, the top SNP*BMI interaction, and BMI into the model. The top SNP for the iteration model is the SNP with the smallest p-value of the interaction coefficient. At each iteration if the interaction coefficient p-value is less than 0.001, we add the top SNP and the top SNP*BMI terms as covariates.

We identified credible sets of Base-eQTL and BMI-eQTL for each eGene by selecting the top SNPs that were conditionally independent and had an original eQTL p-value that was locally significant. Each top SNP marks a discrete credible set that also includes SNPs in LD r^2^ >0.8.

### Interaction Directionality Assignment

For the top SNP in each BMI-eQTL credible set we characterized the direction of the interaction of SNP and BMI into four categories: positive, negative, flipped, and uncertain, using the categorical BMI models. The full decision tree is available in Supplementary Figure 3.

### Transcription Factor Binding Site Analysis

To test if BMI-eQTL disrupt specific transcription factors compared to Base-eQTL we implemented transcription factor binding site (TFBS) affinity analysis using the atSNP package^72^ and the Jaspar2020 motif library. We tested all the variants in each credible set as our TFBS input, annotated to dbSNP151 and computed the affinity scores using ComputeMotifScore() and p-values using ComputePValues(). We assessed the significance of the binding affinity change using an FDR correction of the p-value rank output. We implemented a one-sided binomial test to determine if there is an enrichment for disruption of TFs in the BMI-eQTL. The expected probability *p* is the number of significant rsIDs in a motif X in Base-eQTLs/number of rsIDs tested in motif X in Base. The trials *n* equals the number of rsIDs tested in motif X in BMI-eQTLs and the successes *x* are the number of FDR-significant rsIDs in motif X in the BMI-eQTLs. We implemented an FDR correction to the binomial p-values to correct for the number of motifs tested.

### Cell type deconvolution

We downloaded single cell data across tissues across studies to deconvolute the GTEx bulk data with the most BMI-eQTL. Data was available for cortex^73^, nucleus accumbens^74^, small intestine, transverse colon^75^, and minor salivary gland^76^. We converted all counts data to CPM and used Cibersortx^77^ to construct a signature matrix. We applied that signature matrix to deconvolute the GTEx bulk tissue and obtain estimated cell type fractions for each GTEx sample. We tested if cell type was controlled for in the RNA sequencing quality control, completed with SVA. The estimated cell fractions correlated robustly with surrogate variables (**Sup Fig 4A**) however because we preserved the effect of BMI in SV creation and found BMI correlates with cell fraction there may be residual impacts on discovery. Using the cell fraction and RNA sequencing, we apply Bayesian estimation using bMIND^78^ to estimate cell type-specific gene expression matrices. Of note, when using bMIND only a set number of cell type-specific matrices can be imputed, so at this step we collapse cell types of the same kind (i.e. Excitatory neuron population 1 and 2 are collapsed, Supplementary Table). We then repeat our primary eQTL framework within each cell type to construct cell-type specific eQTL statistics. We determine a lead cell type for each eQTL denoted by the cell type eQTL with the smallest p-value. To determine if there is enrichment for specific cell types amongst lead BMI-eQTL, we completed a binomial test. We tested against the proportion of lead cell type in the Base-eQTL analysis.

### Activity by contact scoring of Interaction-eQTL with ATAC seq

We selected Base- and BMI eGenes credible sets significant in excitatory neurons from deconvolution. We use adapted activity-by-contact (ABC) model using STARE^36^ to score relative regulatory activity of the eQTL in the interaction terms using the R package. The ABC score is calculated using by integrating “activity”, which here is defined by -log10(p-value) of the SNP*BMI interaction coefficient and “contact” which we define using ATAC-sequencing in glutamatergic iPSC neurons along with the distance to eGenes to determine predicted regulatory activity of a SNP toward a specific gene. We assess a baseline contact of ATACseq peaks with no inflammatory markers applied and ATACseq in the context of IL-6, TNF-alpha, and interferon-a. The methods of cell culture can are cited elsewhere^35^. We visualized the distribution of ATAC seq scores across contexts and consulted the literature^79^, and chose to set a threshold requiring the absolute value of the ABC score difference between vehicle and context > 0.02 and ABC score in either vehicle or the context to be > 0.1.

### Creation of BMI-dynamic predictor models

To predict gene expression from SNPs, BMI and SNP*BMI interactions, we use the cv.glinternet() from the glinternet R package^80^. Glinternet employs a Hierarchical Group-Lasso Regularization to select predictive features. The software is intended to learn interactions, and if an interaction coefficient is non-zero then both main effects are included with a non-zero coefficient. We employ a four-fold approach whereby we train the models using a three-fold cross validation and hold out the fourth fold for within-sample validation. We ensured the hold out validation fold has an even distribution of BMI using used StratifiedKFold from the sklearn package in python3.7.3. We treat SNP as a continuous measure, so models can be applied to dosage data. We used the same genotype QC steps described in the eQTL set with a MAF threshold to 5%, which demonstrated reduced overfitting than 1%. For BMI-Interaction model to be significant the cross-fold R2 >0.1 and p < 0.05. Additionally, in the hold-out fold the predicted values must significantly predict the observed values (p-holdout < 0.05).

We tested if the crossfold-R2 significant predicts the 4^th^ fold hold out R2 in each tissue (**Sup Fig 5 C-H**), and found a significant prediction across tissues test with R2 ranging from .314-.765. We find that sample size is vital to learning interactions. In tissues with larger sample sizes, the CV-R2 better predicts the holt out validation R2. We also see the association between CV-R2 and hold out R2 is similar across models that contain just main effects as well as interaction. In smaller tissues, we find that models with interactions have worse association, indicating sample size is paramount in fitting and validating interaction predictor models.

We compare our interaction-aware models to the published GTEx v8 predictor models on PredictDB^81^, which use Predixcan^37^ to predict expression. The published models use elastic net and which account for only the main effects of SNPs using a 1% MAF threshold and have other analysis-specific differences in QC such as PEER versus SVA for latent factor correction. The models made with elastic net in general include more predictors in each model (**Sup Fig 5A**), which may be due to a more lenient MAF filter or features of the differing ML learning approaches.

### BMI-dynamic predictor model application to UK Biobank

The UK Biobank is a cohort of ∼500,000 individuals from the United Kingdom with genotype, lifestyle, health, and anthropometric data. For the UK biobank genotype quality control, we implemented an INFO > 0.8 threshold and filtered relatedness > 0.0625 and subset to European individuals as that is the population used in the published predixcan models. We selected depression and current smoking as psychiatric traits due to epidemiologic associations with BMI^21,82^, the availability of case status and BMI at UKB assessment center visits, and the large sample sizes. To obtain depression case status, we used data field 20002, which is medical history obtained from verbal interview at the assessment center. For current smoking status, we used the lifestyle questionnaire data field 1239 “Current tobacco smoking”, from the touchscreen questionnaire at the assessment center. BMI values were obtained from data field 21001 from the assessment center physical measures.

To calculate predicted expression using the PredictDB models, called GREx, we used the predixcan python package and the elastic net v8 GTEx PredictDB predictor models. To calculate predicted expression for the BMI-interaction glinternet models, we manually multiply each predictor by the beta coefficient output by glinternet in R. The glinternet package scales the input predictors while building the model but returns the unscaled coefficients.

Once predicted expression is calculated, we test the association with psychiatric phenotype using a logistic regression in R using glm() with family = “binomial”. For the trait association analyses we used age, sex, BMI, and PC1-10 as covariates in the predixcan PredictDB models and age, sex, PC1-10 as covariates in the BMI-Interaction model.

We tested across GWAS traits^83,84,85,86,87,88,89,90,91,92,93,94,95,96,97,98^ for evidence of nominal trait association in the identified genes using S-Predixcan^99^. For the 2023 major depressive disorder GWAS, we tested for enrichment of the UK biobank associations in nominal S-predixcan associations using a binomial test.

## Supporting information

Supplementary Results

## Data Availability

All data produced in the present study are available upon reasonable request to the authors

## Acknowlegdements

LMH acknowledges funding from NIMH (R01MH124839, R01MH118278, R01MH125938, RM1MH132648, R01MH136149), NIEHS (R01ES033630), and the Department of Defense (TP220451).

This work was supported in part through the computational and data resources and staff expertise provided by Scientific Computing and Data at the Icahn School of Medicine at Mount Sinai and supported by the Clinical and Translational Science Awards (CTSA) grant UL1TR004419 from the National Center for Advancing Translational Sciences. Research reported in this publication was also supported by the Office of Research Infrastructure of the National Institutes of Health under award number S10OD026880 and S10OD030463. The content is solely the responsibility of the authors and does not necessarily represent the official views of the National Institutes of Health.

## Notes

### Competing Interest Statement

The authors have declared no competing interest.

### Author Declarations

This study used only publicly available data.

