## Supplementary figures and images for "BMI Interacts with the Genome to Regulate Gene Expression Globally, with Emphasis in the Brain and Gut"

### Supplementary Figures

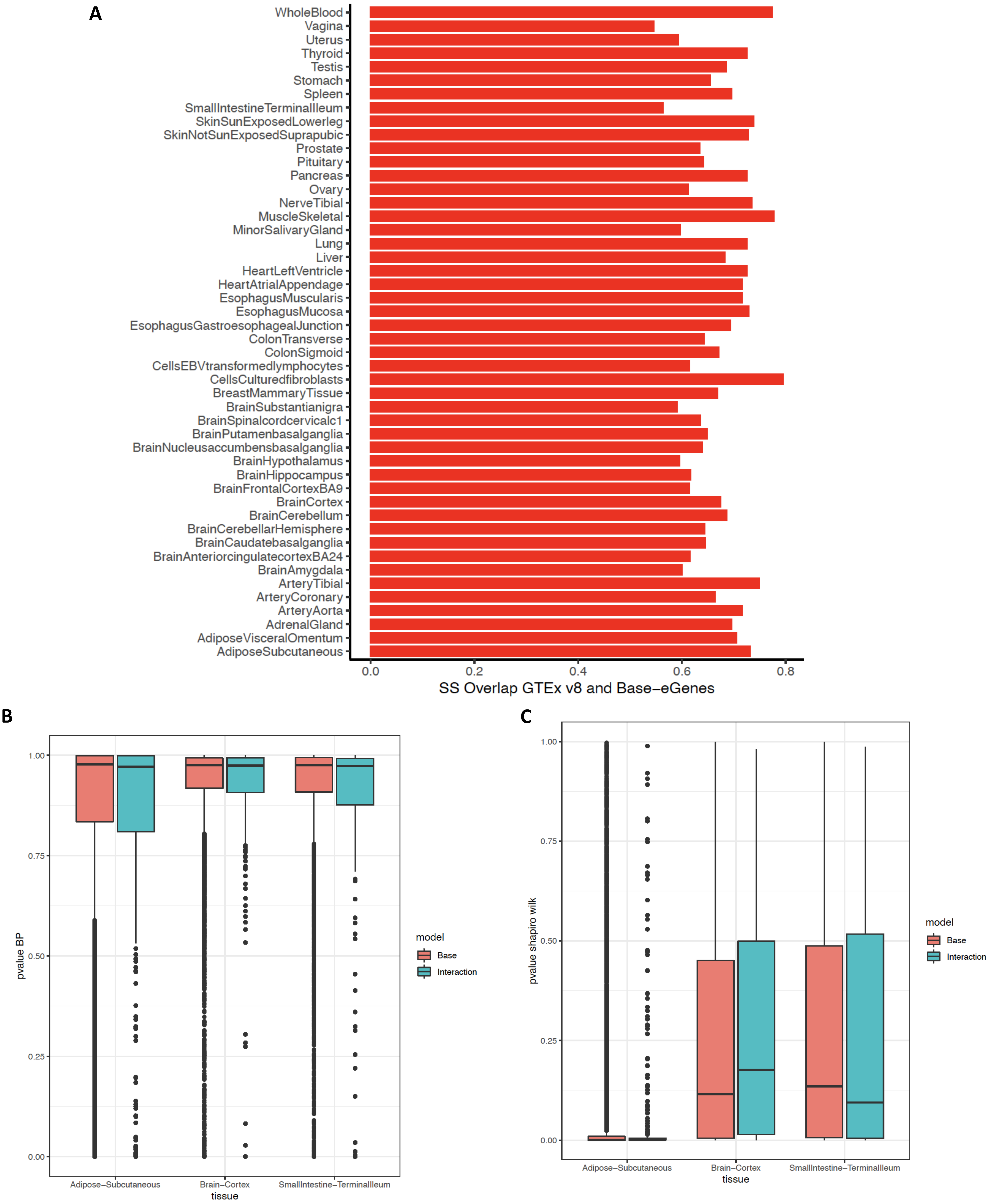

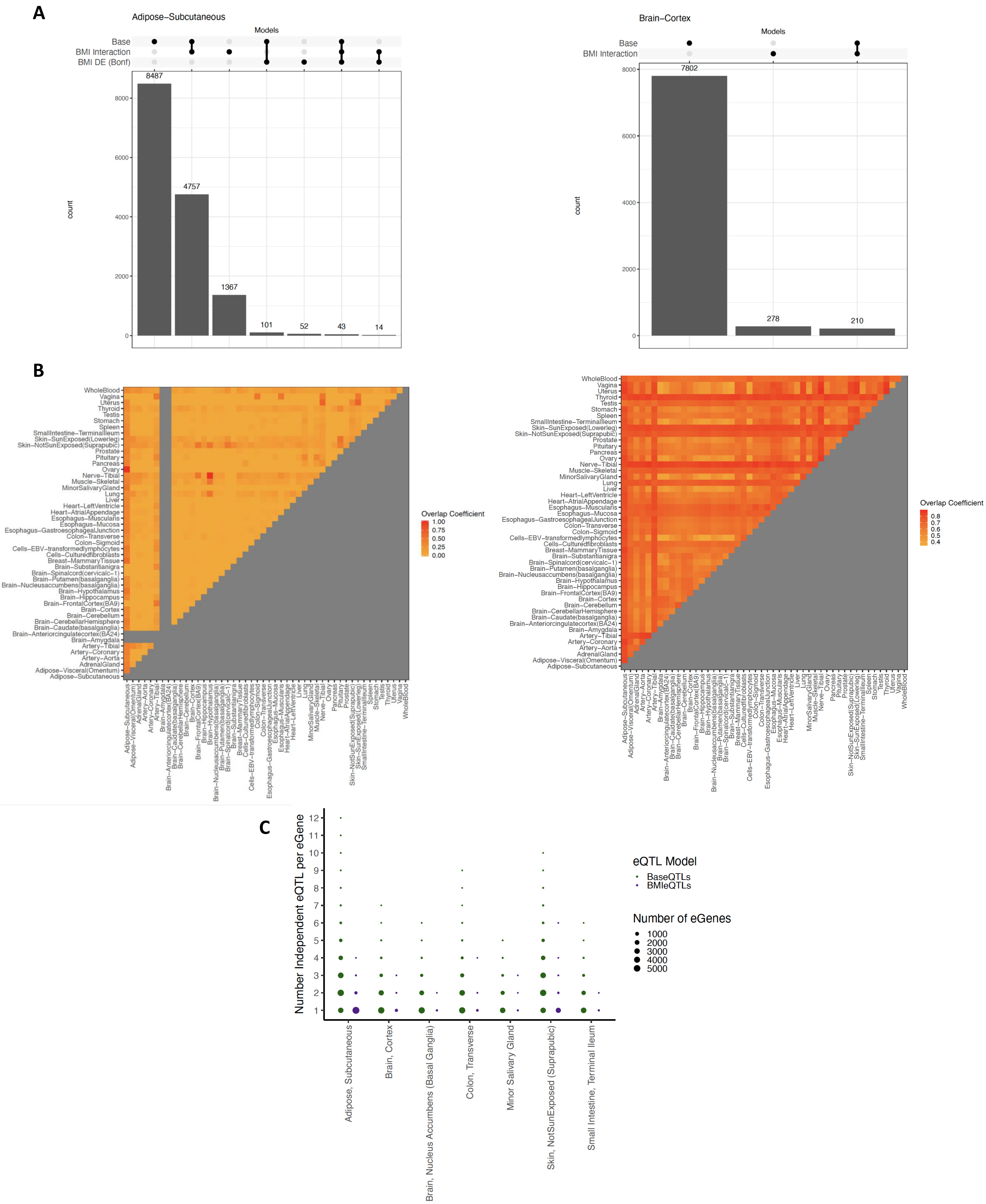

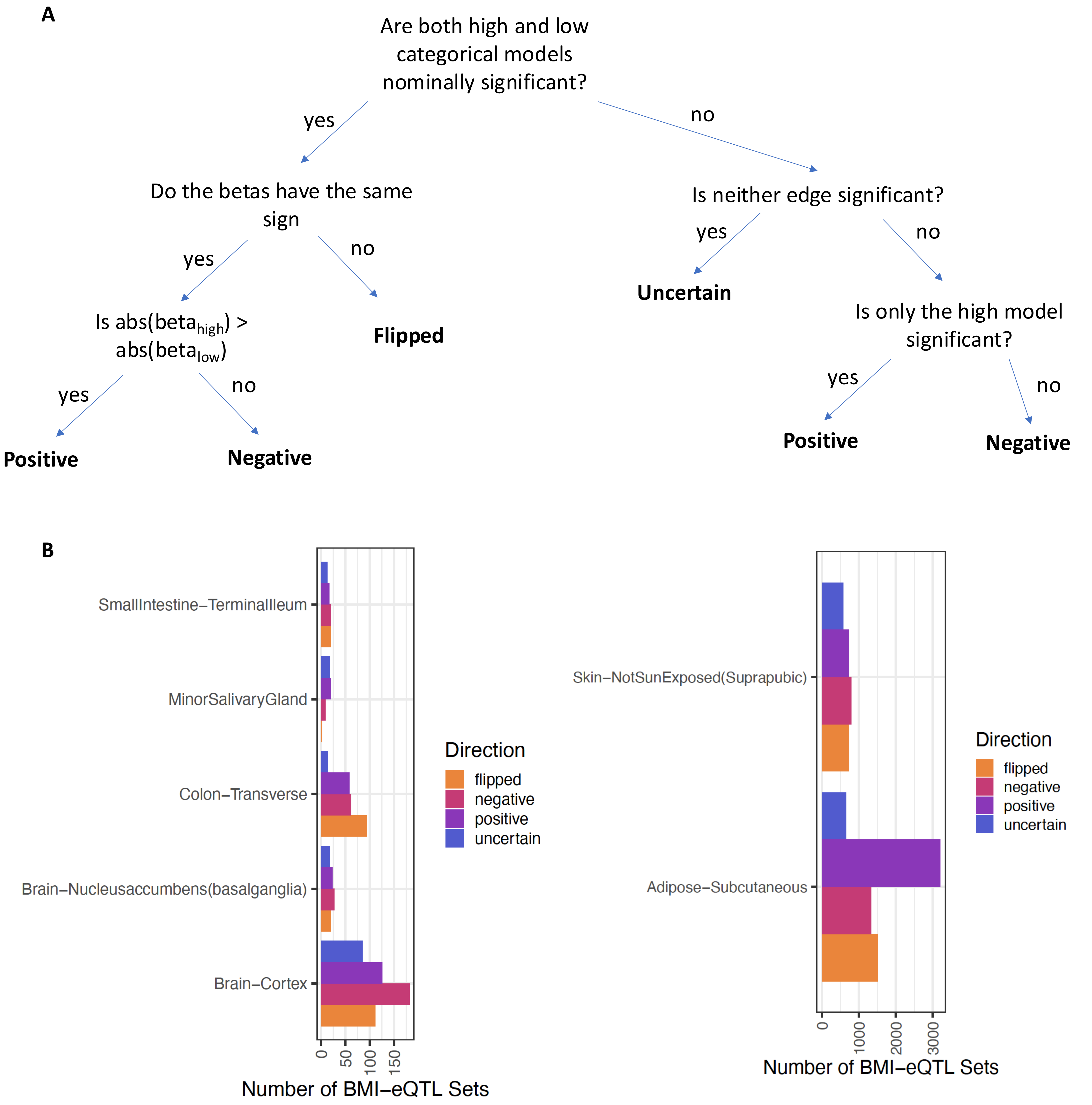

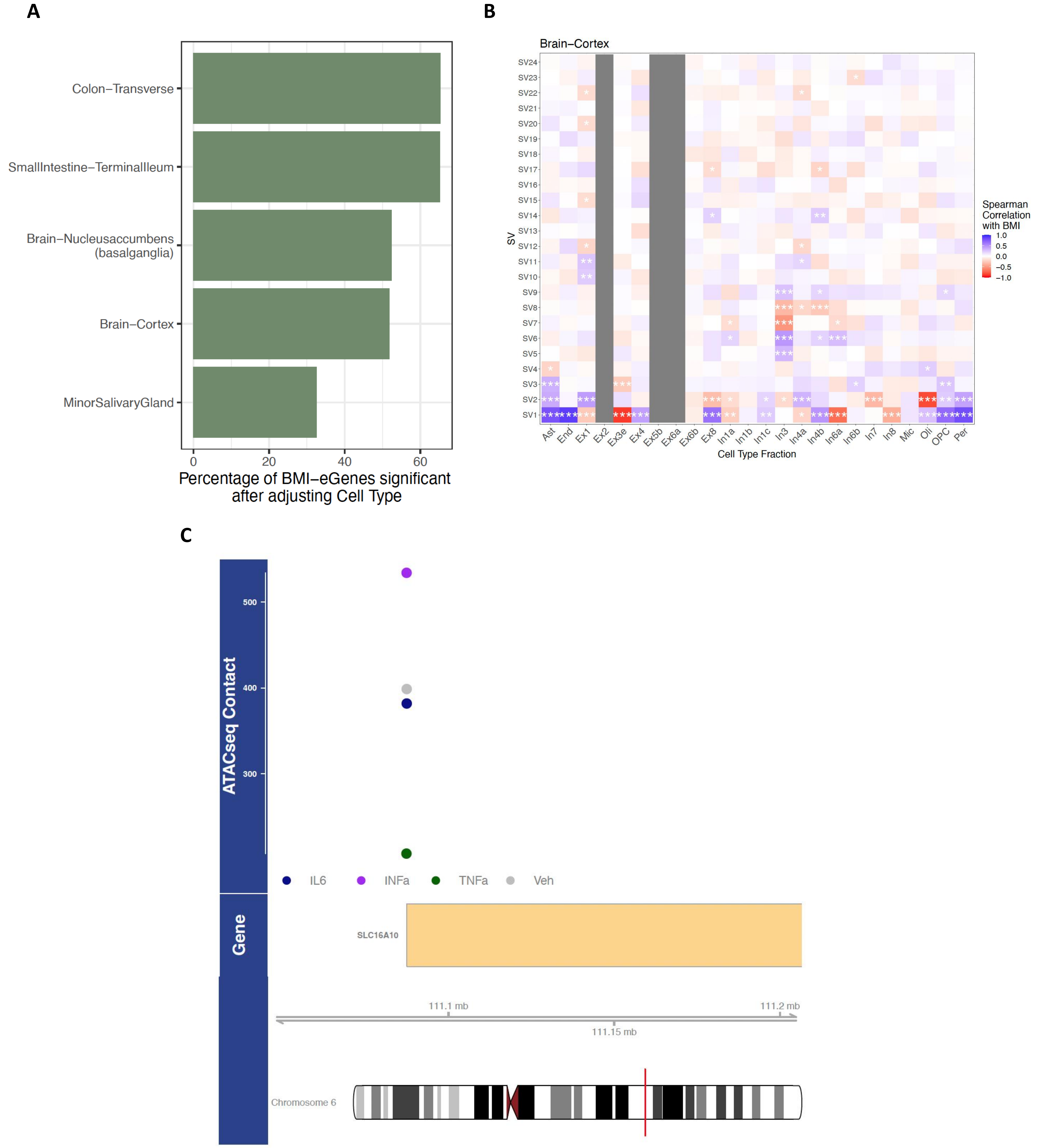

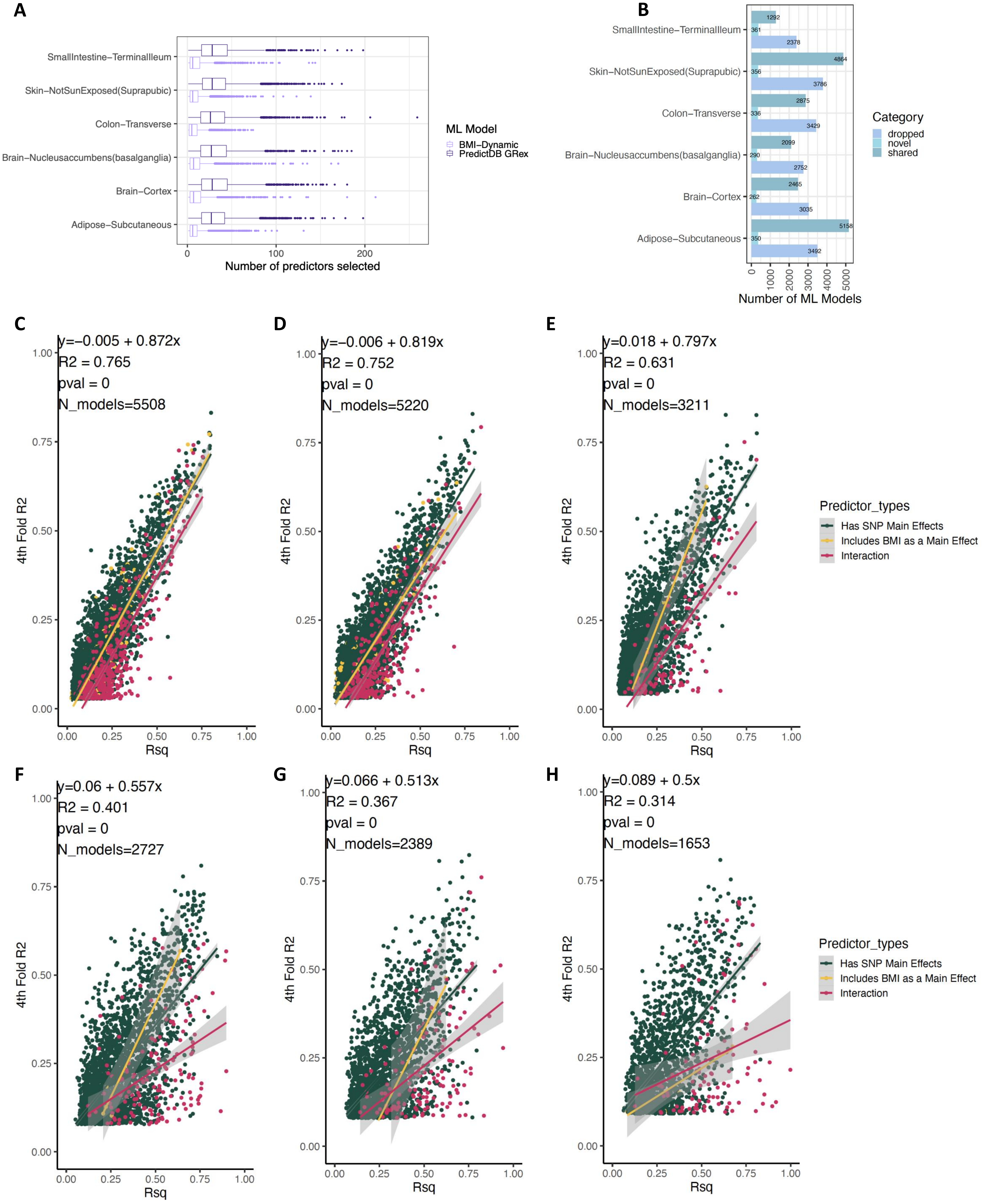

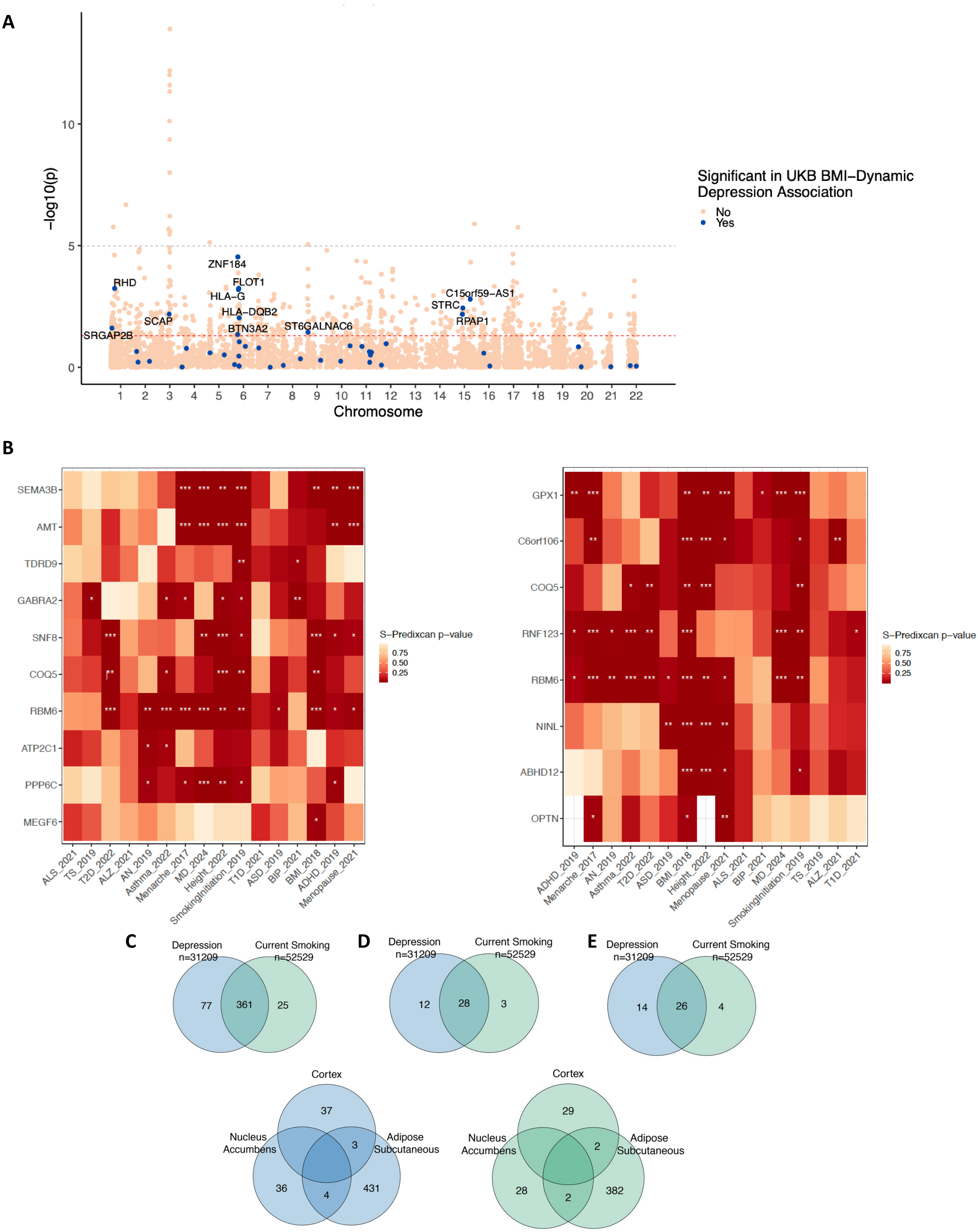
