## Supplementary material for "BMI Interacts with the Genome to Regulate Gene Expression Globally, with Emphasis in the Brain and Gut": Figure Legends

Figure 1. **Identification of Associations of SNP and BMI on Gene Expression in the GTEx post-mortem dataset.** A) Quality control and linear model equations implemented to identify associations, created in Biorender B) Number of locally significant and genome-wide FDR < 0.01 in this Base-eGene analysis compared to GTEx v8 cis-eQTL summary statistics C) Differential expression analysis with colored FDR significant BMI-DE genes, red indicates gene’s expression is negatively associated with BMI, and Blue indicates gene expression is positively associated with BMI, and NS indicates the gene is not FDR significant. D) Number of genes in each tissue that have an FDR < 0.05 significant association with BMI from differential expression analysis, colored according to the primary body system of the tissue. E) Number of BMI-eGenes that are locally significant and FDR < 0.01 across tissues, subset by the number of samples in each tissue where >= 500 samples constitutes large, less than 500 but greater than or equal to 200 constitutes medium, and less than 200 samples constitutes small tissues F) Distribution of significant BMI-eGenes in the Brain: CAU: Caudate, basal ganglia; CNG: Anterior cingulate cortex; PUT: Putamen, basal ganglia; MFC: Cortex; HTH: Hypothalamus; PIT: Pituitary; AMY: Amygdala; HIP: Hippocampus; SN: Substantia Nigra; CB: Cerebellum

Figure 2. **Function and Directionality of BMI-eQTL and eGenes.** A) Within each model (Base-eQTL, BMI-eQTL, and BMI-DE genes) The proportion of genes with a high probability of loss-of-function intolerance (pLI) score and enrichment of high pLI genes compared to the background, * indicates p-binomial < 0.05, ** indicated p-binomial < 0.01, *** indicates p-binomial < 0.001 B) FDR Significant gene sets enriched in tissues not including Adipose and tissues with N-BMI-eGenes < 20 C-F) Examples of directionality of eQTL showing the data in the significant interaction model equation (top) and the categorical models (bottom), which are used to define the direction. Of note, BMI categories were separated into quintiles where m1, m2, and m3 represent the three middle quintiles. Red dashed lines indicate the threshold for low and high BMI quintiles within tissue. Light blue points indicate individuals who are homozygous reference, dark blue points are heterozygous alternate, and purple are homozygous alternate C) Example of a positive BMI-eQTL: chr12_55900153_A_G_b38 regulating *IKZF4* in Transverse Colon D) Example of a negative BMI-eQTL: chr4_5281524_A_G_b38 regulating *CRMP1* in Cortex E) Example of a flipped BMI-eQTL: chr9_111930711_G_A_b38 regulating *LPAR1* in Adipose, Subcutaneous F) Example of uncertain interaction: chr17_32888312_G_T_b38 regulating *PSMD11* in Minor Salivary Gland G) The directionality category that is the most numerous within tissue

Figure 3. **Parsing the Impact of Sequence and Cell Type on BMI-dynamic Functions.** A) Enrichment for predicted transcription factor affinity binding changed in BMI-eQTL credible sets compared to Base-eQTL credible sets B) Methodologic summary of cell type deconvolution and cell-type specific eQTL calling in GTEx bulk mRNA sequencing data created in Biorender C) Spearman Correlation of predicted cell type proportion and BMI within each tissue examined, labeled are those with a significant p< 0.05 correlation with BMI D) FDR significant gene set enrichment in genes significant after cell types are corrected and within cell-type in small intestine and transverse colon, intersection size > 1 E) Cell types with significant enrichment amongst BMI-eQTL lead SNPs in credible sets compared to Base-eQTL

Figure 4. **Functional validation of Cortex BMI-eQTL in iPSC Glutamatergic Neurons.** A) Lead cell types of top SNPs in Base-eGenes (*Left*), and BMI-eGenes (*Right*), excitatory neurons are starred to indicate these credible sets are prioritized for modeling in iPSC system. B) Genes with evidence of activity by contact in at least one context (ABC >= 0.1) and evidence of a shift in activity compared to vehicle (|ABC-Context – ABC-Vehicle| >= 0.02) C) Amongst SNPs with an ABC-shift, SNPs that have significant transcription factor affinity binding score D) The locus of the SNP rs6927697 with the BMI-eQTL statistic for gene *SLC16A10*, ABC score, FDR-significant MZF1 binding prediction on chromosome 6.

**Figure 5. Model Creation to Predict Gene Expression using BMI, SNPs, and BMI-SNP Interactions** A) Graphical representation of models built with main and interactive effects created in Biorender B) The number of BMI-dynamic models created using glinternet compared to the number of models in the published PredictDB GREx data C) The difference in cross fold R-squared from BMI-dynamic models compared to the current published PredictDB GREx models, colored by the types of predictors selected and annotated with the p-value of the t-test. Of note, the glinternet package forces the main effects to be included if the interaction is selected. D) In the BMI-dynamic models, the number of models created that select BMI as a main effect for each tissue, separated by if BMI is the sole predictor in that gene’s model, if SNPs are present as well, or if a BMI-SNP interaction is present

Figure 6. **Gene Discovery in Psychiatric Traits using Machine Learning** A) Graphical Representation of association analysis completed in UK Biobank to identify gene-tissues associated with psychiatric traits created in Biorender B) The number of genes with predicted expression significantly associated with each trait in the UK biobank using both the published PredictDB GREx models and the BMI-dynamic models C) Gene set enrichment of genes significant in each tissue-trait association compared to the background of genes expressed in that tissue D) For genes not identified in the PredictDB-PrediXcan, the distribution of predictor types in each significant model E) S-predixcan association statistics from 16 GWAS of complex traits for genes significant in both the BMI-eGene analysis and the UKB association analysis are subset in cortex. X axes are labeled with trait abbreviation and year of the GWAS. Trait abbreviations are as follows: AN_2019=anorexia nervosa; MD_2024=Major depression, 2024; ALS_2021=amyotrophic lateral sclerosis; BIP_2021=bipolar disorder- all; ADHD_2019=Attention-deficit/hyperactivity disorder; BMI_2018=Body mass index; T2D_2022=Type 2 diabetes; TS_2019=Tourette Syndrome; ALZ_2021=Alzheimer; Menarche_2017=Age at Menarche; Menopause_2021=Age at Menopause
