## Supplementary Results for "BMI Interacts with the Genome to Regulate Gene Expression Globally, with Emphasis in the Brain and Gut"

**Supplemental Results**

To determine whether BMI-eQTL replicate across data sets, we repeated the eQTL analysis in the CommonMind Consortium (CMC) dorsolateral prefrontal cortex (DLPFC-CMC). We tested the overlap of GTEx Brain-Cortex genome-wide FDR < 0.01 significant eGenes and CMC genome-wide FDR < 0.05 eGenes and found an SS overlap coefficient of 0.82 for the Base analysis and 0.39 for the interaction. We identified two BMI-eGenes, *ELAPOR2 and MPP4* that were FDR < 0.01 in both sets. 194/488 (40%) of Cortex BMI-eGenes were both FDR < 0.01 in GTEx and FDR < 0.05 in CMC. The SNP pi1 overlap in 0.55 and 0.076 in base and interaction, respectively. Replication may be lower for BMI-eQTL as age, BMI, and genome coverage ranges for the two studies.

**Supplemental Figure legends**

**Figure S1 Comparison of eQTL and eGene sets.** A) Szymkiewicz–Simpson (SS) overlap coefficient between significant cis-eGenes in GTEx v8 publication and Base-eGene analysis from this study. B) Breusch-Pagan (BP) test for heteroskedasticity p-value for linear models in the BaseQTL and BMIeQTL model equations C) Shapiro-wilk test for normality p-value for linear models in the BaseQTL and BMIeQTL model equations.

**Figure S2 eGene overlap and complexity across models and tissues.** A) Overlap of eGenes across Base-eGenes, BMI-eGenes, and BMI-DE genes in adipose subcutaneous (left) and brain-cortex (right) B) Szymkiewicz–Simpson (SS) overlap Coefficient measure of eGenes across tissues in the BMI-eGene model (Left) and Base-eGene model (Right) C) The number of independent eQTL signals per eGene following forward stepwise conditional analysis green on the left shows Base-eQTL model and purple on the right shows BMI-eQTL model

**Figure S3 Interaction Direction Characterization with Categorical BMI Models.** A) Decision tree that accepts the high and low BMI quintile eQTL statistics to sort BMI-eQTL into positive, negative, flipped, and uncertain. Positive BMI-eQTL have greater absolute activity at high BMI, negative have greater absolute activity at low BMI, flipped BMI-eQTL have flipped direction of effect, and uncertain BMI-eQTL have no significant association at either edge. B) The number of BMI-eQTL with each direction in each tissue, separated by sample size of the GTEx tissue

**Figure S4 Cell type and context-specific effects in BMI-eQTL Discovery.** A) The percentage of BMI-eGenes what are eigenMT-FDR significant after correcting for Cibersortx-predicted cell type proportion in the BMI-eQTL equation B) The correlation of surrogate variables used to residualize the RNA sequencing expression matrix in quality control and cell type proportion estimated by cibersortX C) Context-specific ATACseq contact at the promoter region of *SLC16A10* for IL-6, TNFa, and INFa

**Figure S5 Machine Learning Model Selection and Performance.** A)The Number of Genes with significant CV and hold out R2 in glinternet BMI-interaction models subset by if the gene is present in the published PredictDB-Predixcan elastic net GTEx v8 models. Novel indicates models are not in published, shared indicates a model is made in both sets, and dropped indicates a model is made in the published models but absent from the glinternet BMI-Interaction models B) The number of significant models made using glinternet BMI-Interaction that only contain SNP main effects and no BMI interactions C-H) Cross-fold validation R2 on the x-axis compared to hold-out R2 on the y-axis for all significant models. The statistics from the equation: *hold-outR2 ~ CV-R2 printed* are shown in text in C) Adipose, subcutaneous D) Skin, not sun exposed E) Colon, transverse F) Brain, cortex G) Brain, nucleus accumbens H) Small intestine, terminal ileum

**Figure S6 Properties of significant gene-trait associations in UK Biobank ML Model Application** A) The –log10(p-values) of the S-predixcan output in the Nucleus Accumbens from the most recent 2024 MD GWAS, colored if the gene is also significant in the Nucleus Accumbens BMI-Dynamic gene expression associations in the UK Biobank Depression phenotype B) S-Predixcan nucleus accumbens statistics for a number of well-powered traits with BMI associations, showing genes that *Left)* are significant in the UK Biobank PredictDB GREx predicted gene expression *Right)* Genes significant in the BMI-Dynamic association and only have SNPs in the glinternet model C-E) In the UK Biobank BMI-dynamic trait association, the number of genes shared and unique between depression and smoking in C)Adipose, subcutaneous D) Brain Cortex E) Brain nucleus accumbens F-G) Within each UK Biobank trait, the number of BMI-dynamic association genes shared across tissues in F) Depression and G) Current Smoking
